# Variants in Interferon Lambda are Associated with Very Early Onset Inflammatory Bowel Disease

**DOI:** 10.1101/2022.03.17.22271929

**Authors:** Jodie D. Ouahed, Achille Broggi, Abigail Glick, Robin T. Haring, Michael Field, Daniel Chinnapen, Michael J. Grey, Wayne Lencer, Steven J. Steiner, Evida Dennis-Heyward, Noah Shroyer, Zachary Criss, Shih-Ching Lin, Xi-Lei Zeng, Sue E. Crawford, Mary K. Estes, Jay R. Thiagarajah, Amalia Capilla, Whitney Scoon, Gustavo Mostoslavsky, Jason Spence, Christoph Klein, Aleixo M. Muise, Bruce Horwitz, Ivan Zanoni, Scott B. Snapper

## Abstract

Inflammatory bowel diseases (IBD) are chronic inflammatory disorders of the intestine that affect children and adults. The etiology is multifactorial with the contribution of genetic, immune, microbial, and environmental risk factors with a substantial involvement of host:microbial cross-talk in the lumen. Whether and how antiviral responses contribute to IBD pathogenesis is under intense investigation. Here, we identified two unrelated patients diagnosed with very early onset IBD (VEOIBD) with rare variants in type III Interferons (IFNs), also known as interferon lambdas (IFNλs), a group of IFNs that play key roles in the protection against enteric viruses. The first patient was found to have homozygous variants in both *IFNL2* and *IFNL3*, the genes encoding, IFN-λ2 and IFN-λ3, respectively. The second patient was found to have inherited compound heterozygous variants in *IFNL3*, with one of the two alleles bearing the same variant as the first patient. Functional analyses revealed that the proteins coded for by these variant IFN-λ genes exhibited defects in the induction of IFN signaling. More detailed assessment of the variants identified in Patient 1 demonstrated defects in their ability to bind to IFN-λ receptor 1 (IFNLR1), as well as their ability to induce heterodimerization of IFNLR1 and IL10RB, which together compromise the functional receptor for IFN-λ (IFNLR). These patient-encoded IFN-λ variants also exhibit defects in the ability to induce robust downstream IFN-stimulated genes (ISGs) in patient-derived intestinal organoids. All in all, we demonstrate that VEOIBD is associated with variants in IFN-λs and defective induction of downstream IFN signaling.

## Introduction

Inflammatory bowel diseases (IBDs), encompassing Crohn’s disease and ulcerative colitis, are chronic inflammatory conditions affecting the intestines. IBD is now considered a global disease, with prevalence exceeding 0.3% in North America^1^. While most patients are diagnosed in adulthood, IBD also affects infants and young children. Diagnosis before 6 years of age, referred to as very early onset IBD (VEOIBD)^2^, represents the group with the fastest rising incidence of IBD^3^. Overall, the etiology of IBD is felt to be multifactorial with contributions of genetic, immune, microbial, and environmental risk factors.

Large adult-based Genome Wide Association Studies (GWAS) have identified nearly 250 loci that confer significant risk for IBD susceptibility^4-6^. It has been hypothesized that the genetic contribution of VEOIBD is larger than that in patients diagnosed at older ages; currently over 75 monogenic causes of VEOIBD have been identified with many reflecting underlying primary immune deficiencies^7-11^. These advances have not only significantly expanded our understanding of the pathophysiology of IBD but importantly, they also enable personalized and often curative interventions.

Crosstalk between the microbiome and the genetically susceptible host ^12-18^ is a central factor in the development of IBD. However, recent data has also implicated the *virome* in mucosal homeostasis^19-25^, and interestingly, to date, none of the currently identified monogenic causes of VEOIBD directly implicate defects in viral recognition or clearance. Interferons (IFNs) are most known for their inherent anti-viral activities and protect against enteric viral infections^26-34^. IFNs are divided into three classes: type I IFNs (including IFN-α and IFN-β), type II IFN (IFN-γ) and the more recently identified type III IFNs or IFN-λs (including IFN-λ1-4 in humans^35-37^). Murine models have suggested potential anti-inflammatory functions for type I IFNs within the intestine^38^, and based on this, type I IFNs were assessed for therapeutic efficacy in IBD^39,40^. However, these trials failed to demonstrate therapeutic efficacy; in fact type I IFNs exacerbated intestinal inflammation in some cases^41^. Recent murine studies revealed potential insight into these failures including opposing effects of type I IFN signaling on individual cell types^42^. Also, type I IFN-dependent ISGylation of proteins produces reactive oxygen species and augments colonic inflammation^43^.

IFN-λs were initially believed to have functions redundant with those of type I IFNs. However, more recent research by our group has established unique characteristics of IFN-λs. IFN-λs can induce signaling in both murine and human neutrophils^44^. IFN-λ suppresses the tissue damaging functions of neutrophils and the specific loss of IFN-λ signaling in neutrophils exacerbates DSS-induced colitis in mice ^44^. This may have important therapeutic implications as the exogenous administration of IFN-λ reduces intestinal inflammation, whereas IFN-λ blockade enhances intestinal inflammation^44^. Also, it has recently been shown in mice that IFN-λ participates in the generation of amphiregulin for regeneration of intestinal epithelium^45^. While previous studies have demonstrated that expression of IFN-λ and its receptor, the interferon lambda receptor (IFNLR), are upregulated within the colon of patients with IBD, the effects of IFN-λ signaling during IBD development are incompletely understood^46,47^. Nonetheless, IFN-λ demonstrates features which support a potential role in protection against intestinal inflammation^48-51^. First, IFN-λ belongs to the IL-10 cytokine family and IFN-λs are structurally more similar to the anti-inflammatory cytokine IL-10 than type I IFNs. The IFNLR comprises a heterodimer consisting of IL10RB (shared by IL-10, IL-22 and other cytokines) and IFNLR1, which confers IFN-λ recognition. Pertaining specifically to impaired mucosal homeostasis, the absence of IL10RB signaling in innate immune cells results in life-threatening VEOIBD, with a major role for IL-10 signaling residing in phagocytes^52^. Secondly, IFN-λs activate only a subset of the genes that are activated by type I IFNs, with the different signaling capacities of the two IFN groups explained, at least in part, by their activation of different JAK-STAT members^44,49,53^ that have been found to be associated to IBD through GWAS studies^54-56^. Finally, the cellular targets of IFN-λs are much more limited and defined, compared to those of type I IFNs, because the IFNLR1 is almost exclusively expressed on epithelial cells and neutrophils, suggesting tissue-specific functionality^48,57^.

Here, we describe two patients from two unrelated families that have rare variants in IFN-λ in association with VEOIBD. Functional studies demonstrate that the variants lead to defects in binding to the heterodimeric IFNLR and induction of IFN signaling.

## Results

### Clinical presentation of two unrelated patients with VEOIBD

The first patient, P1, presented with bloody diarrhea in the first months of life. He was formally diagnosed with indeterminate colitis at 1 year of age following endoscopic (**Figure 1A**) and histologic findings consistent with VEOIBD. He also experienced large joint arthritis and intermittent fevers. His disease was well managed on the immune modulator 6-mercaptopurine. The medication was discontinued by 5 years of age but symptoms recurred several years later. Reinitiation of 6-mercaptopurine with adequate levels failed to control disease.

**Figure 1:**
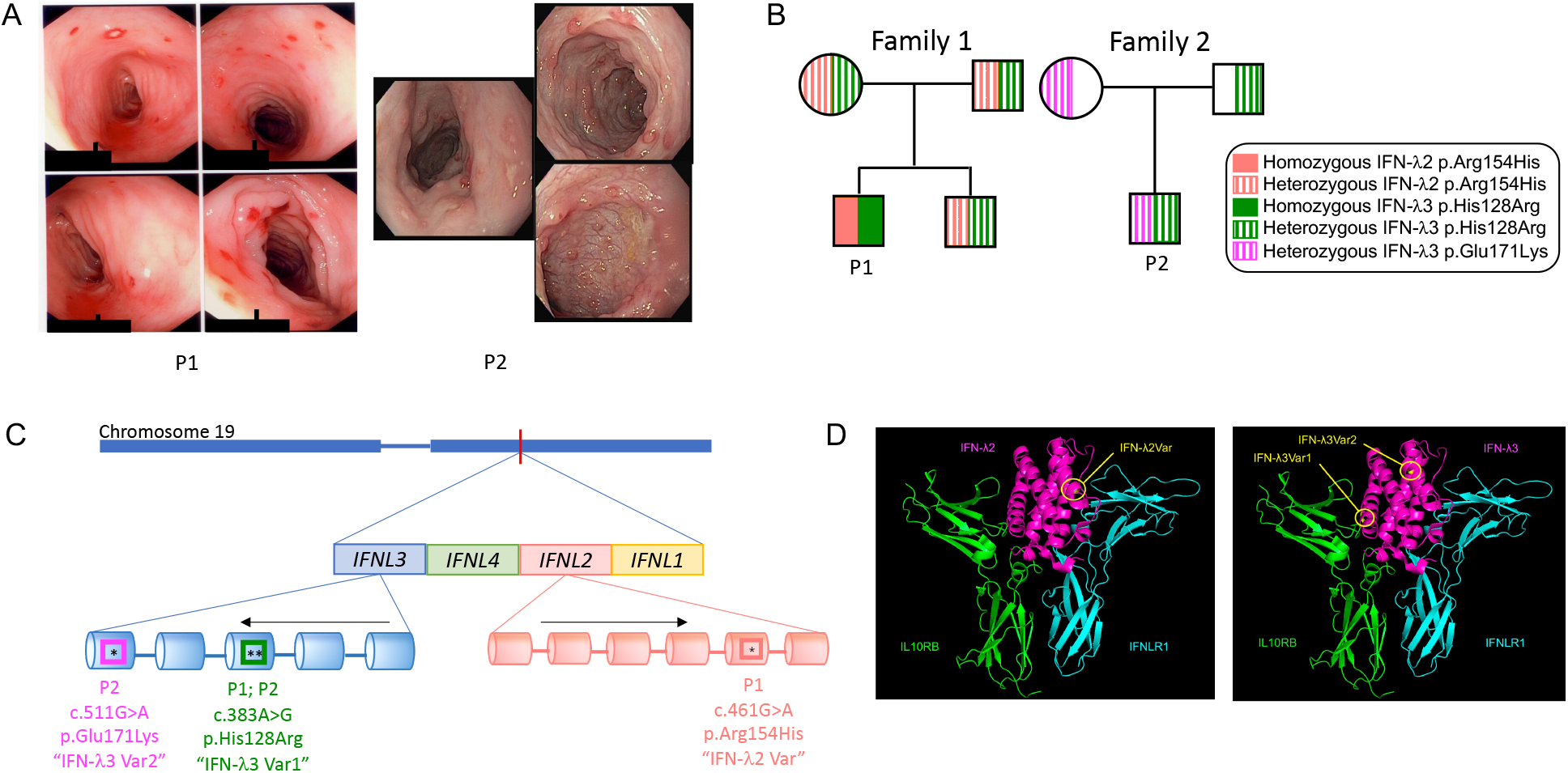
Variants in IFN-λs identified in two unrelated patients with VEOIBD: **(A)** Colonoscopy pictures of P1 and P2, revealing loss of vascular markings, and numerous discrete aphthous ulcerations throughout. **(B)** Pedigrees of 2 unrelated patients with VEOIBD and mutations in IFN-λ. P1 inherited 2 homozygous (solid color) mutations: One in IFN-λ2 (peach) the other in IFN-λ3 (green). His healthy parents and brother are heterozygous (hatched lines) for each variant. P2 inherited a compound heterozygous mutation in IFN-λ3, including the same variant in IFN-λ3 as does P1 (green), and another (fuchsia). **(C)** Schematic representation of IFN-λ2 (orange) and IFN-λ3 (blue) genes within chromosome 19 (blue bar on top), illustrating 6 exons (cylinders) in IFN-λ2 and 5 exons in IFN-λ3. Black arrows reflect direction of transcription. The asterisk demonstrates the location of each mutation. Representation is not to scale. **(D)** 3-dimensional crystalized structure of WT IFN-λ2 and estimated location IFN-λ2Var, extrapolated from WT IFN-λ3 using PyMOL software given that there is no available crystal structure of IFN-λ2 (left panel). 3-dimensional crystalized structure of WT IFN-λ3 and location of IFN-λ3Var1 and IFN-λ3Var2 with respect to its heterodimeric receptor (IL10RB and IFNLR1) using PyMOL software (right panel)

The second patient, P2, presented with bloody stools also in the first months of life. By 1 year of age he was found to have endoscopic (**Figure 1A**) features consistent with VEOIBD. Parents refused medical therapeutic intervention. By 5 years of age, parents report that his condition spontaneously improved with complete resolution of his bloody stools.

### Variants in interferon lambda are associated with VEOIBD

To identify potential monogenic causes of VEOIBD, whole genome sequencing (WGS) was performed on P1 and his healthy parents. No rare variants in genes known to be responsible for VEOIBD were identified. Whole exome sequencing (WES) was performed on P2 and his healthy parents and similarly did not reveal rare variants in genes associated with VEOIBD.

To identify potential novel monogenic causes of P1 and P2’s VEOIBD, we evaluated the presence of rare variants in genes not currently known to be related to VEOIBD. In P1, we identified homozygous missense mutations in both IFN-λ2 and IFN-λ3, which are arranged in a cluster of IFN-λ genes on chromosome 19. The mutation in IFN-λ2 is c.461G>A; p.Arg154His, referred to as “IFN-λ2Var” for the remainder of this manuscript. The mutation in IFN-λ3 is c.383A>G; p.His128Arg, referred to as “IFN-λ3Var1” for the remainder of this manuscript (**Figure 1B and 1C**). The IFN-λ2Var had a minor allele frequency (MAF) of <0.1% in both 1000 Genomes Project^58^and ExAC^59^. It was reported a single time in a homozygous fashion in ExAC. IFN-λ3Var1 was reported <0.1% in 1000 genomes and ExAC and had never been reported in a homozygous fashion. Both variants were confirmed by Sanger sequencing and present in a homozygous fashion in patient P1 and inherited in a heterozygous fashion in his healthy parents and healthy brother (*not shown*).

In P2, we identified biallelic rare missense variants in IFN-λ3 (**Figure 1B and 1C**); specifically, c.383A>G; His128Arg (i.e., IFN-λ3Var1 identified in P1), as well as c.511G>A; p.Glu171Lys, referred to as “IFN-λ3Var2” for the remainder of this manuscript. The latter missense mutation had a minor allele frequency < 0.1% and was reported 9 times in a homozygous recessive fashion in ExAC. These mutations were confirmed by Sanger sequencing (*not shown*) and found to be present in a compound heterozygous fashion in the patient as illustrated in the pedigree in **Figure 1B**.

Mapping of the mutation on a published crystal structure of IFN-λ3 complexed with the heterodimeric receptor, consisting of IFNLR1 and IL10RB,^60^ revealed that IFN-λ3Var1 was situated in a site close to the residues critical for IL10RB binding (**Figure 1D, right panel**). IFN-λ3Var2 is located closer to the binding site of IFNLR1 (**Figure 1D, right panel**). Given the great similarity between IFN-λ3 and IFN-λ2 (which share 96% amino acidic sequence identity)^61^, we mapped the location of IFN-λ2Var on the IFN-λ3-IL10RB-IFNLR1 crystal complex. IFN-λ2Var appeared situated near the binding site of IFNLR1 (**Figure 1D, left panel**).

### Patient-encoded variants in IFN-λ2 and IFN-λ3 result in reduced activation of interferon signaling

We sought to determine whether the rare patient-encoded variants in IFN-λ2/3 resulted in defective downstream signaling. Upon binding of IFN-λ to the IFNLR, activation of JAK1 and JAK2 or TYK2 results in phosphorylation of STAT1, STAT2 and recruitment of IRF9, and formation of the ISGF3 complex. ISGF3 binds IFN sensitive response elements (ISREs) and induces a variety of downstream IFN-stimulated genes (ISGs) which confer antiviral activity. To determine the capacity of the patient-encoded variants to induce signaling downstream of the IFNLR, we employed a luciferase reporter assay driven by the ISRE promoter and assessed relative luciferase activity as a proxy for induction of downstream ISGs in 293T cells^49^. Cells were treated overnight with a range of doses of either wild type (WT) or patient-encoded variant IFN-λ2 or IFN-λ3. Patient-encoded variants IFN-λ2Var, IFN-λ3Var1 and IFN-λ3Var2 had significantly reduced signaling as compared to WT IFN-λ2 or IFN-λ3 (**Figure 2A-D, Supp Figure 1A-E**). Importantly, the shared variant IFN-λ3Var1 resulted in negligible downstream induction of signaling (**Figure 2C-D, Supp Figure 1B-C**).

**Figure 2:**
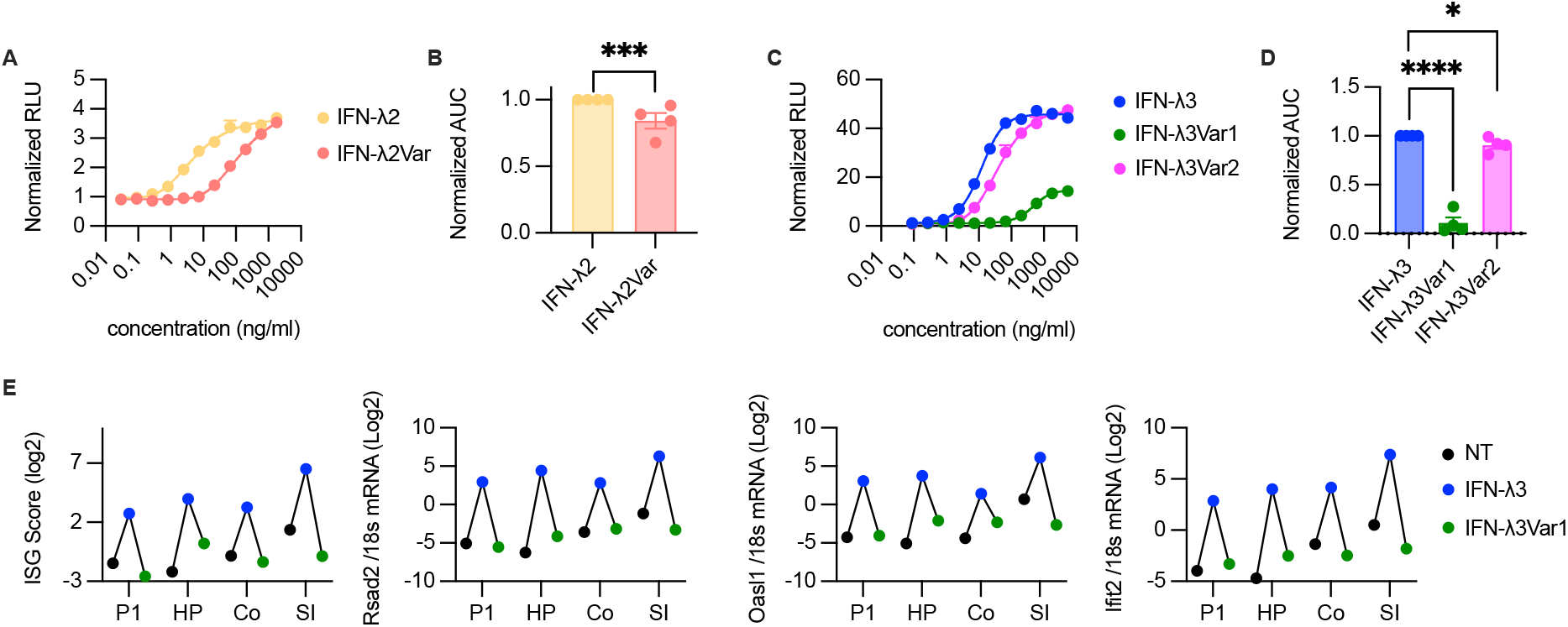
Patient-encoded IFN-λ2/3 variants have reduced activation of ISGs: **(A)** Dose-response curve of downstream ISG induction following stimulation with either IFN-λ2 or IFN-λ2Var protein. Normalized relative luciferase units (RLU) shown. Data was normalized to blank wells (no protein). **(B)** AUC comparison between IFN-λ2 and IFN-λ2Var. **(C)** Dose response curve of downstream ISG induction following stimulation with either WT or patient-encoded IFN-λ3 proteins (IFN-λ3Var1 and IFN-λ3Var2). Data was normalized to blank wells (no protein). **(D)** AUC comparison among IFN-λ3, IFN-λ3Var1 and IFN-λ3Var2. **(E)** Induction of ISGs following stimulation of patient-derived tHIOs with either wild-type IFN-λ3 (blue dots), patient-encoded IFN-λ3Var1 (green dots), or no treatment (black dots). P1 = Patient; HP = healthy parent; Co = colonic enteroids of unrelated healthy control; SI = small intestinal enteroids of unrelated healthy control. (A-D) 3 independent experiments are depicted; each data point is an independent experiment. RLU were normalized to blank wells (no protein) for each independent experiment. Statistical significance for (B, D) was assessed by one sample t (B) and Wilcoxon (D) test.

### Patient 1- and parent-derived intestinal organoids respond to wild-type but not Patient 1-encoded variant IFN-λ2 and IFN-λ3

We have shown that both P1 and P2 harbor variants in IFN-λ2 and IFN-λ3 that display defective signaling in a reporter cell line. To assess whether these defects are conserved in patient-derived intestinal epithelial cells, we focused on PI and differentiated, as previously described^62^, human intestinal organoids (HIOs) from induced pluripotent stem cells (iPSCs) derived from his PBMCs as well as from his parents. These *in vitro* HIOs were then transplanted in immunodeficient mice, subsequently the epithelium of the transplanted HIOs (tHIOs) were extracted and expanded to facilitate further study. As controls, we also evaluated human intestinal enteroids derived from duodenal biopsies (SI) or colon biopsies (CO) of healthy controls. We then determined whether P1 or P1 parent tHIOs responded to WT or mutant IFN-λs by measuring by qPCR the induction of a set of 3 ISGs, whose mean expression was quantitated as an “ISG score” as previously described^63^. Our data not only confirmed that IFN-λ2 and IFN-λ3 variants are hypoactive (**Figure 2E, Supp Figure 1F**), but also demonstrated that P1 tHIOs responded normally to WT IFN-λ3 (**Figure 2E)**.

### Patient 1-encoded mutations in IFN-λ result in altered binding and dimerization of the IFN-λ heterodimeric receptor

To interrogate the basis for the deficient downstream signaling of patient-encoded variants, we sought to determine in P1 whether the IFN-λ2 and IFN-λ3 variants resulted in either altered receptor binding and/or impaired dimerization of the heterodimeric IFN-λ receptor. As IFN-λ directly binds IFNLR1, and does not bind directly to the IL10RB component of the heterodimeric receptor^60^, we focused our attention on the capacity of WT IFN-λs and variant IFN-λs to bind IFNLR1.

We employed a bio-layer interferometry (BLI)-based receptor binding affinity assay to assess binding of WT IFN-λ2 and P1-encoded IFN-λ2Var to IFNLR1. Reduced binding affinity was detected in the patient-encoded protein (K_D_ 2.5×10^−6^ M, versus 2.0×10^−7^ M) **(Figure 3A)**. The receptor binding affinity assay was also performed with WT IFN-λ3 and the shared patient-encoded IFN-λ3Var1. Compared to WT IFN-λ3, IFN-λ3Var1 had reduced binding affinity to IFNLR1 (K_D_ 1.0×10^−6^ M, versus 5.9×10^−8^ M) **(Figure 3B)**.

**Figure 3:**
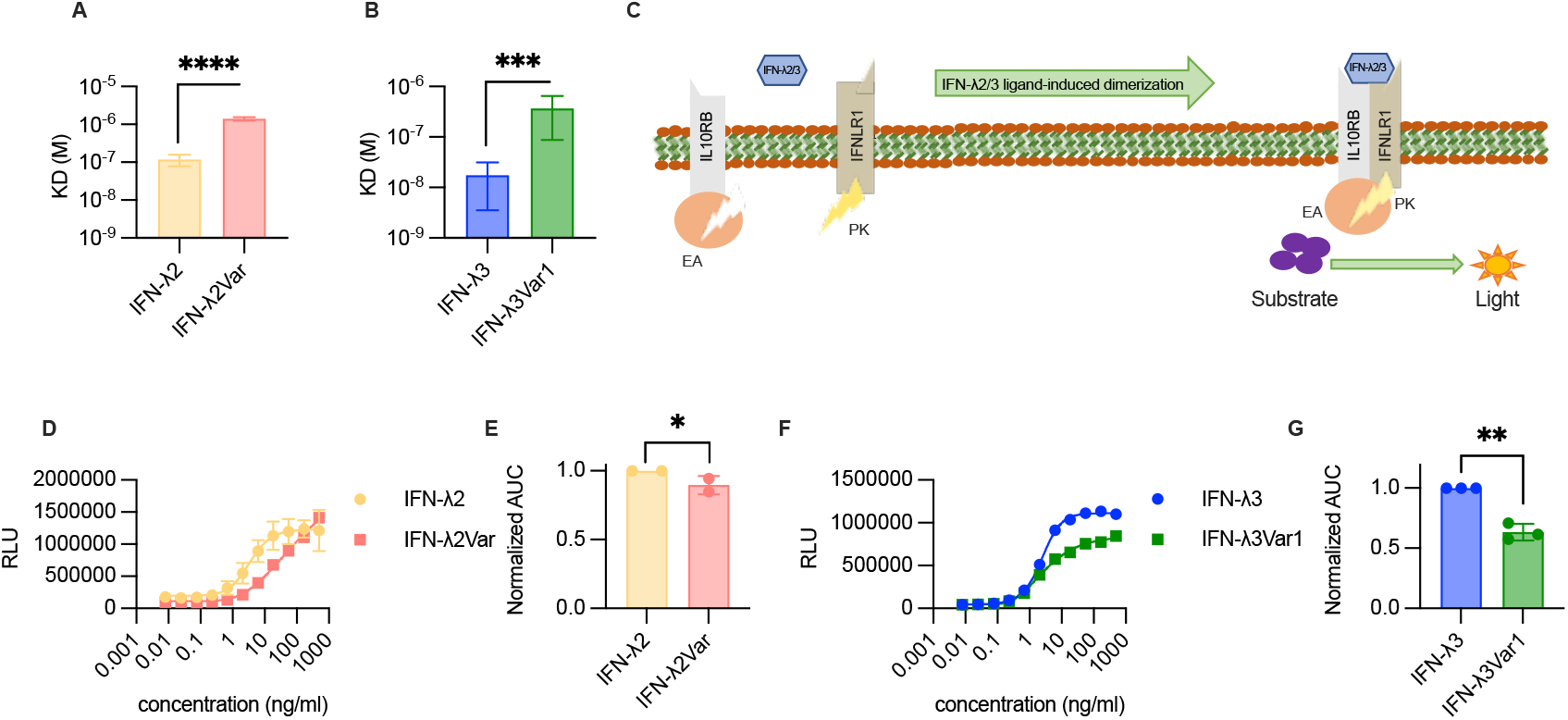
Patient-encoded IFN-λ variants have reduced IFNLR1 binding and elicit reduced IFN-λ receptor dimerization. **(A)** K_D_ calculated via receptor binding affinity assay (bio-layer interferometry) of wild type IFN-λ2 and patient-encoded IFN-λ2Var to IFNLR1. Mean and SEM of 6 replicates is depicted. **(B)** K_D_ calculated via receptor binding affinity assay (bio-layer interferometry) of wild type IFN-λ3 and patient-encoded variant protein IFN-λ3Var1 to IFNLR1. Mean and SEM of 6 technical replicates is depicted. **(C)** Schematization of dimerization assay. **(D)** RLUs reflecting dimerization of IFNLR1 and IL10RB following binding with either IFN-λ2 or IFN-λ2Var. **(E)** AUC comparison for dimerization assays of the heterodimeric receptor upon binding of IFN-λ2 or IFN-λ2Var. **(F)** RLUs reflecting dimerization of IFNLR1 and IL10RB following binding with either IFN-λ3 or IFN-λ3Var1. **(G)** AUCs comparison for dimerization assays of the heterodimeric receptor upon binding of IFN-λ3 or IFN-λ3Var1. (D-G) Data representing two independent experiments (D, E) or three independent experiments (F, G), each being the average of two replicates. Significance of differences in (E, G) was assessed using a one sample t and Wilcoxon test.

Similar to other IL-10 family cytokines, IFN-λs are recognized first by the specific subunit of their dimeric receptor, i.e., IFNLR1. Upon binding to IFNLR1, they recruit and bind to IL10RB. Receptor dimerization is thus the first step to induce signaling. As described above, IFN-λ3Var1 and IFN-λ2Var had reduced binding affinity to IFNLR1. We then sought to assess whether P1-associated variants resulted in impaired dimerization of the IFNLR heterodimeric receptor. We used an IFNLR1/IL10RB dimerization assay (PathHunter eXpress, Europhins DiscoverX Products) in which each component of the heterodimeric receptor is linked to a portion of an enzyme with β-galactosidase activity. Namely IFNLR1 is linked to an enzyme acceptor (EA) and IL10RB is linked to a ProLink (PK) enzyme. Each component individually lacks β-galactosidase activity (**Figure 3C**); however, upon cytokine-induced receptor dimerization, a functional β-galactosidase is formed that can hydrolyze a chemiluminescent substrate. As expected from its IFNLR1 binding defect, patient-encoded IFN-λ2Var induced defective receptor dimerization (**Figure 3D-E, Supp Figure 1G)**. Patient derived IFN-λ3Var1 induced a severe reduced dimerization of the receptor (**Figure 3F-G, Supp Figure 1H**), consistent with the localization of the mutation in the IL10RB binding region (**Figure 1D, right panel**) and the severe defect in signaling induced by IFN-λ3Var1 (**Figure 2C-D, Supp Figure 1B-C**).

## Discussion

While an abundance of genetic and functional studies has supported dysregulation of intestinal bacteria in association with IBD, there has been enhanced focus on the contribution of other microbes, and notably viruses, in disease pathogenesis^19-25^. IBD risk polymorphisms have been described (e.g., FUT2 and FUT3) that may regulate the replication of enteric viruses^25,64^. Nonetheless, to our knowledge, no monogenic cause of IBD associated with a specific defect in enteric virus infection has been reported to date.

Here we present two unrelated patients with VEOIBD who inherited rare recessive missense variants in IFN-λs. P1 inherited rare homozygous recessive mutations in both IFN-λ2 and IFN-λ3, while P2 inherited a compound heterozygous mutation in IFN-λ3, and of interest, one of the variants was shared with P1. We provide functional evidence of deleterious downstream signaling of each of the patient-encoded mutations (IFN-λ2Var; IFN-λ3Var1; IFN-λVar2) as evidenced by significantly attenuated dose response curves of generation of downstream ISG activation using a luciferase reporter assay, with the most pronounced deficit identified in the shared variant IFN-λ3Var1. We focused on the patient-encoded variants in P1 and determined that IFN-λ2Var and IFN-λ3Var1 elicit defective induction of ISGs in both patient- and control-tHIOs and reduced binding affinity to the IFNLR1 component of the heterodimeric IFNLR, as evidenced by a higher K_D_ than in their respective WT proteins. Moreover, we found that IFN-λ2Var and IFN-λ3Var1 result in reduced dimerization of the components (IFNLR1 and IL10RB) of the IFNLR.

Relevance of IFN-λ in regulating intestinal mucosal homeostasis has been clearly demonstrated in mice^44,65^. Mice unable to signal through IFNLR1 have enhanced mucosal inflammation and mice devoid of both IFN-λ2 and IFN-λ3 (IFN-λ1 is a pseudogene in mice) have defective mucosal anti-viral responses^65^. Murine norovirus, reovirus, and influenza A virus have enhanced intestinal replication in mice lacking either IFNLR1 or both IFN-λ2 and IFN-λ3 ^65^. Moreover, exogenous administration of IFN-λ can suppress intestinal inflammation in wild-type mice following DSS-induced colitis^44^. These data are especially supportive of the intestinal disease identified in P1.

It is intriguing that both patients improved significantly over time, as evidenced by the lack of flares and discontinuation of medical therapy by 5 years of age in P1 and the spontaneous resolution of bloody stools in P2 by 5 years of age without any medical intervention. There are several hypotheses that may explain this atypical clinical progression of their VEOIBD. One possibility is that since humans have 4 IFN-λ cytokines, the members of the type III IFN family are differentially regulated over time. IFN-λ4 is a pseudogene in a large part of the population^66^, while IFN-λ1, IFN-λ2 and IFN-λ3 are highly homologous (81%-96% at the aminoacid level).^67^ It is thus possible that in our affected patients (with defective IFN-λ2 and/or IFN-λ3) the expression of one of the other intestinal IFN-λs (i.e., IFN-λ1) may have compensated for the lack of function. Notably, a recent study by our group has shown that there is compartmentalization of IFN-λ expression in the respiratory tract of SARS-CoV-2 infected patients depending on disease severity^68^. Specifially, IFN-λ1 and IFN-λ3 drive protective ISGs in the upper airways of mildly ill patients, whereas critically ill patients express IFN-λ2 and have low ISG expression. Moreover, we demonstrated that different cell types (i.e., epithelial cells vs. phaogcytes), preferentially produce specific members of the type III IFN family. The nature of the IFNs produced, the cell origin, and the transcriptional programs initiated by specific members of the IFN-λ family will need to be taken into consideration to explain how these IFNs impact health and disease in the human intestine. In particular, while ISGs that are induced by type III IFNs are expressed in subsets of mature enterocytes in the small intestine at homeostasis^69^, a comprehensive understanding of the specific cellular, developmental (e.g., peri-natal versus post-natal), and geographic (e.g., small versus large intestine) landscape of type III IFN expression in the human intestine is lacking.

While we have identified rare IFN-λ variants that are functionally deficient in our described patients, it remains to be determined definitively whether these functional deficits relate directly to impaired mucosal homeostasis and development of VEOIBD. These possibilities are not mutually exclusive and require further investigation needs to be pursued.

## Data Availability

All data produced in the present study are available upon reasonable request to the authors

## Acknowledgements

SBS is supported by NIH grant P30DK03485, RC2DK122532, R01DK115217, the Wolpow Family Chair in IBD Treatment and Research, the Translational Investigator Service at Boston Children’s Hospital, and the Children’s Rare Disease Cohort (CRDC) Study. IZ is supported by NIH grant 1R01AI121066, 1R01DK115217, 1R01AI165505 and contract no. 75N93019C00044, Lloyd J. Old STAR Program CRI3888, and holds an Investigators in the Pathogenesis of Infectious Disease Award from the Burroughs Wellcome Fund. AB is supported by: the excellence initiative of Aix Marseille Université-A*Midex, a French “investissements d’Avenir” program: AMX-20-CE-01; the FRM amorçage de jeunes equipes grant AJE202010012468 and the ANR-JCJC grant “INTERMICI” ANR-21-CE15-0022. JO is supported by NIH grant K08 DK122133-01A1, the Boston Children’s Hospital Translational Research Program Mentored Translational Investigator Service Award. RTH is supported by the AGEM International Student Research Fellowship Grant Next generation sequencing was supported by Merck Research Labs. X-LZ, S-CL, SEC and MKE are supported by NIH grants DK P30DK056338, P01 AI 057788 and R01 AI 080656.

## Materials and Methods

### Patient identification

Patients were referred to us at Boston Children’s Hospital from offsite centers through the very early onset IBD consortium (www.veoibd.org). Patients and parents included in this study provided informed consent and assent where applicable. Consent was obtained as per our research protocols as was publication of de-identified information in this work, through our Institutional Review Board (protocol# P00000529). De-identified information was stored on RedCap through the Boston Children’s Hospital (BCH) Pediatric Gastrointestinal Data Registry and BioRepository.

### Whole Exome and Whole Genome Sequencing

Whole Exome Sequencing (WES) was performed in collaboration with Merck Research Labs using an Agilent v5 Sureselect capture kit and Illumina 2500 sequencing technology as previously described^70^. For each sample, paired end reads (2×100 bp) were obtained, processed and mapped to the human reference genome (hg19). We used the BWA mem algorithm (version 0.7.12)^71^ for alignment of the sequence reads. The HaplotypeCaller algorithm of GATK version 3.4 was applied for variant calling, as recommended in the best practice pipeline^72^. KGG-seq v.08 was used for annotation of identified variants^73^ and in house scripts were applied for filtering based on family pedigree and local dataset of variants detected in previous sequencing projects.

Whole genome sequencing was performed as described^74^ using the HiSeq platform (Illumina) with the Burrows–Wheeler aligner, and Picard was used for basic alignment and sequence quality control. The same capture kits and strategies were employed for the genome sequencing as described above for whole exome sequencing.

Next Generation Sequencing data was analyzed by a biostatistician (MF) in conjunction with trained gastroenterologists (JO, SBS). Proband variants were filtered by rare, loss-of-function (LOF) or missense variants inherited according to standard Mendelian inheritance patterns. Rare variants were those with minor allele frequency (MAF) <1% for homozygous, hemizygous, and de novo mutations and <5% for compound heterozygous mutations according to ExAC^59^ and the 1000 genomes project^58^.

### Protein purification of IFN-λ2, IFN-λ3 and patient encoded IFN-λ variants

Proteins were isolated and purified by two different methods. Both methods yielded comparable results.

#### First method

IFN-λ proteins were expressed in the methylotrophic yeast *Pichia pastoris*. Genes corresponding to IFN-λ2 (wild type and variant R154->H) and IFN-λ3 (wild type and variant 1 H128->R), were synthesized by Integrated DNA Technologies (Coralville, IA) and cloned into PICZa-A with a C-terminal hexahistidine tag and secretion signal from EasySelect™ *Pichia* Expression Kit (ThermoFisher, MA). Following linearization and integration into *P. pastoris* strain GS115, the highest expressing clones were isolated and used for large scale expression. Protein was expressed over the course of 2-3 days at 1.5 mL scale at 30°C in BMMY media (1% yeast extract, 2% peptone, 100mM potassium phosphate pH 6.0, 1.34% YNB, 0.04 mg% biotin and 0.5% methanol) and expression induced by addition of methanol to 1%. Media was harvested and secreted proteins purified by Ni-NTA (GoldBio, MO) after pH adjustment to 7.7. Further purification was done by SP-HiTrap cation exchange column (Cytiva, MA) and buffer exchanged into PBS by Amicon centrifugal filtration (SigmaMillipore, MA). Final purity of constructs was estimated >97% based Coomassie gel and Western blot. Protein concentration was assessed by UV absorbance at 280nm.

#### Second method

IFN-λ2 and IFN-λ3 and patient-derived mutant constructs were PCR amplified using Phusion GC Master Mix (New England Biolabs, Ipswich, MA) and subcloned into pcDNA4-myc-his-B expression vector (Thermo Fisher, Waltham, MA) using EcoRI and AgeI restriction sites to yield a C-terminal His6 tag. All constructs were verified by Sanger sequencing. Plasmids were propagated in NEB5alpha cells (New England Biolabs) and purified using PureLink Expi Endotoxin-Free Mega Plasmid Purification Kit (Thermo Fisher). His-tagged constructs were expressed by transient transfection in Expi293 suspension cells according to manufacturer’s instructions (Thermo Fisher). At 4-5 days post-transfection, the conditioned media was collected and adjusted to 20 mM HEPES, 300 mM NaCl, 20 mM imidazole, and 1X Complete protease inhibitor. His-tagged proteins were bound to cobalt resin (GoldBio, St Louis MO) equilibrated in wash buffer (20 mM HEPES pH 7.4, 300 mM NaCl, 20 mM imidazole), washed with 2 column volumes (CV) of wash buffer, and eluted with 8x 0.5 CV fractions of elution buffer (20 mM HEPES pH 7.4, 150 mM NaCl, 250 mM imidazole). Elution fractions were assayed by SDS-PAGE and Coomassie staining, pooled, and buffer exchanged into phosphate buffered saline using PD10 desalting columns. The concentrations were estimated by absorbance at 280 nm using calculated extinction coefficients of 12865 M^-1^ cm^-1^ for IFN-λ2 and 11375 M^-1^ cm^-1^ for IFN-λ3 (ProtParam). Samples were analyzed by gel filtration chromatography for monodispersity and differential scanning fluorimetry for thermal stability.

### Assessment of ISG activation in response to IFN-λ2, IFN-λ3 and patient-encoded variants

293T cells expressing Luciferase under the control of an ISRE promoter^49^ were plated at a concentration of 400,000 cells/ml of 293T medium (RPMI medium 1640 1X 500 ml, 1% glutamax, 10% filtered fetal bovine serum (FBS), 1% Pen-strep, 1% non-essential amino acids, 1% sodium pyruvate (ThermoFisher Scientific)) using 100ul per well on a 96-well flat bottom plate in the morning. After adhering to the well, the cells were stimulated with different concentrations of either WT purified protein or patient-encoded proteins. Following overnight stimulation at 37°C, luciferase activity was quantified the following morning using Bright-Glo luciferase assay system (Promega, Madison, WI) on a Tecan plate reader (Männedorf, Switzerland). For assessment of IFN-λ2 and IFN-λ2Var, experiments were performed in triplicates, at concentrations between 0-1800 ng/ml. For assessment of IFN-λ3, IFN-λ3Var1 and IFN-λVar2, experiments were performed > 3 times, with consistent results at concentrations between 0-200 ng/ml.

### Assessment of binding of IFN-λ2, IFN-λ3 and patient-encoded variants to heterodimeric IFN-λ receptor via bio-layer interferometry (BLI)

Binding of IFN-λ3, IFN-λ2 and patient derived variants to IFNLR1 was evaluated via biolayer interferometry (BLI) using an Octet RED384 (Fortebio). With BLI, one molecule is immobilized to a Dip and Read Biosensor and binding to a second molecule is measured as a shift in the light interference pattern measured in real time. We first loaded an anti-human IgG Fc Capture (AHC) Biosensor (Fortebio) with 10 μg/ml of recombinant IFNLR1-Fc chimeric protein (R&D Systems) for 120 sec in the presence of assay buffer (PBS 0,5% Tween-20). The biosensor was then transferred to a different well containing IFN-λ2, IFN-λ3 or patient-derived variants. A range of concentration was assessed (2μM – 1μM – 0.5μM – 0.125 μM – 0.0625 μM) and association to IFNLR1 was measured over 300s. Following the association step, the biosensor was transferred in a well containing assay buffer alone and dissociation was measured over 600s. Association curves were normalized based on the signal of the last 2 seconds of the loading step. Dissociation constants (K_D_) were calculated using the Octet Data Analysis HT Software.

### Assessment of dimerization of heterodimeric IFN-λ receptor

As per the protocol for PathHunter eXpress IFNLR1/IL10RB dimerization Assay protocol (Eurofins DiscoverX Products, San Francisco, CA), cells provided in the kit were thawed as per protocol. Increasing concentrations of IFN-λ2 and IFNλ-3 and patient-encoded variants were added to a final concentration of 0 to 500ng/ml. Cells were stimulated for 6 hours at 37°C prior to adding 110ul of PathHunter Flash Detection reagents to each well, as per protocol. Cells were then incubated at room temperature in the dark for 1hr and luciferase was read using a Tecan plate reader (Männedorf, Switzerland).

### Generation of patient and healthy control human intestinal organoids

Induced Pluripotent Stem Cells (iPSCs) were derived from patients’ and healthy parents’ Peripheral Blood Mononuclear Cells (PBMCs) as previously described^75^. Human intestinal organoids (HIOs) were generated from patient’s and control (parents’) iPSCs as outlined in McCracken et al^62^. To obtain epithelial only organoid lines, HIOs were transplanted into the kidney capsule of immunocompromised *Nod/Scid/Il-2rα*^*-/-*^ mice and extracted after 8 weeks. The epitehlium of transplants were then isolated and propagated as tHIOs, as previously described^76^. tHIOs were maintained in media adapted from Watson et al^76^, consisting of: Advanced DMEM/F12, glutaMAX-I (2 mM), HEPES (10 mM), Wnt, Rspondin, and noggin (WRN) condition media, B27 supplement (1x), N2 supplement (1x), N-acetylcysteine (1 mM), mouse recombinant EGF (50 ng/ml), A-83-01 (500 nM), SB202190 (10 μM), nicotinamide (10 mM), [Leu15]-Gastrin I (10 nM), Y27632 (10 μM), Rspondin condition media (1x), primocin (100 μg/ml), normacin (100 μg/ml).

### Generation of healthy control small intestine and colonic biopsy-derived enteroids

Colonic and duodenal biopsy samples were obtained from routine diagnostic endoscopy under Boston Children’s Hospital IRB protocol P00027983 and cultured with methods modified from Sato et al^77^. Briefly, crypts were dissociated from colonic and duodenal biopsy samples obtained from age-matched (<3yrs) healthy control individuals. Isolated crypts were suspended in Growth Factor Reduced Phenol Red Free Matrigel (Corning, Corning, NY) and plated as 50µL domes in a tissue culture–treated 24-well plates (Thermo Fisher Scientific, Waltham, MA) with growth factor (Wnt3, R-spondin, Noggin) supplemented media. Enteroids were passaged as previously described. Briefly, enteroids were passaged by removal of Matrigel with Cell Recovery Solution (Corning) for 30 min at 4°C. After incubation organoids were mechanically dissociated and replated in Matrigel every 4 days.

### Quantification and Statistical Analysis

To compare two groups unpaired t-test were performed. When data did not meet the normality assumption, significance was assessed by Mann-Whitney test. To establish the appropriate test, normal distribution and variance similarity were assessed with the D’Agostino-Pearson omnibus normality test. Dose response curves were fitted with a Richard’s five parameters dose response curve. EC50 and area under the curve (AUC) were calculated with GraphPad PRISM software. All statistical analyses were two-sided and performed using GraphPad PRISM software software and are indicated in figure legends. Throughout the paper significant is defined as follows: ns, not significant (p > 0.05); *p < 0.05, **p < 0.01, ***p < 0.001, and ****p < 0.0001.

## References

1. Ng, S. C. et al. Worldwide incidence and prevalence of inflammatory bowel disease in the 21st century: a systematic review of population-based studies. Lancet 390, 2769–2778, doi:10.1016/S0140-6736(17)32448-0 (2018).

2. Muise, A. M., Snapper, S. B. & Kugathasan, S. The age of gene discovery in very early onset inflammatory bowel disease. Gastroenterology 143, 285–288, doi:10.1053/j.gastro.2012.06.025 (2012).

3. Benchimol, E. I. et al. Trends in Epidemiology of Pediatric Inflammatory Bowel Disease in Canada: Distributed Network Analysis of Multiple Population-Based Provincial Health Administrative Databases. Am J Gastroenterol, doi:10.1038/ajg.2017.97 (2017).

4. Jostins, L. et al. Host-microbe interactions have shaped the genetic architecture of inflammatory bowel disease. Nature 491, 119–124, doi:10.1038/nature11582 (2012).

5. Liu, J. Z. et al. Association analyses identify 38 susceptibility loci for inflammatory bowel disease and highlight shared genetic risk across populations. Nat Genet 47, 979–986, doi:10.1038/ng.3359 (2015).

6. de Lange, K. M. et al. Genome-wide association study implicates immune activation of multiple integrin genes in inflammatory bowel disease. Nat Genet, doi:10.1038/ng.3760 (2017).

7. Pazmandi, J., Kalinichenko, A., Ardy, R. C. & Boztug, K. Early-onset inflammatory bowel disease as a model disease to identify key regulators of immune homeostasis mechanisms. Immunol Rev 287, 162–185, doi:10.1111/imr.12726 (2019).

8. Ouahed, J. et al. Very Early Onset Inflammatory Bowel Disease: A Clinical Approach With a Focus on the Role of Genetics and Underlying Immune Deficiencies. Inflammatory bowel diseases 26, 820–842, doi:10.1093/ibd/izz259 (2020).

9. Kelsen, J. R. et al. Exome sequencing analysis reveals variants in primary immunodeficiency genes in patients with very early onset inflammatory bowel disease. Gastroenterology 149, 1415–1424, doi:10.1053/j.gastro.2015.07.006 (2015).

10. Uhlig, H. H. et al. The Diagnostic Approach to Monogenic Very Early Onset Inflammatory Bowel Disease. Gastroenterology 147, 990–1007 e1003, doi:10.1053/j.gastro.2014.07.023 (2014).

11. Uhlig, H. H. et al. Clinical Genomics for the Diagnosis of Monogenic Forms of Inflammatory Bowel Disease: A Position Paper From the Paediatric IBD Porto Group of European Society of Paediatric Gastroenterology, Hepatology and Nutrition. J Pediatr Gastroenterol Nutr 72, 456–473, doi:10.1097/MPG.0000000000003017 (2021).

12. Kamada, N., Seo, S. U., Chen, G. Y. & Nunez, G. Role of the gut microbiota in immunity and inflammatory disease. Nat Rev Immunol 13, 321–335, doi:10.1038/nri3430 (2013).

13. Nell, S., Suerbaum, S. & Josenhans, C. The impact of the microbiota on the pathogenesis of IBD: lessons from mouse infection models. Nat Rev Microbiol 8, 564–577, doi:10.1038/nrmicro2403 (2010).

14. Frank, D. N. et al. Molecular-phylogenetic characterization of microbial community imbalances in human inflammatory bowel diseases. Proc Natl Acad Sci U S A 104, 13780–13785, doi:10.1073/pnas.0706625104 (2007).

15. Gophna, U., Sommerfeld, K., Gophna, S., Doolittle, W. F. & Veldhuyzen van Zanten, S.J. Differences between tissue-associated intestinal microfloras of patients with Crohn’s disease and ulcerative colitis. J Clin Microbiol 44, 4136–4141, doi:10.1128/JCM.01004-06 (2006).

16. Sonnenburg, J. L. & Backhed, F. Diet-microbiota interactions as moderators of human metabolism. Nature 535, 56–64, doi:10.1038/nature18846 (2016).

17. Rooks, M. G. & Garrett, W. S. Gut microbiota, metabolites and host immunity. Nat Rev Immunol 16, 341–352, doi:10.1038/nri.2016.42 (2016).

18. Lee, W. J. & Hase, K. Gut microbiota-generated metabolites in animal health and disease. Nat Chem Biol 10, 416–424, doi:10.1038/nchembio.1535 (2014).

19. Yang, J. Y. et al. Enteric Viruses Ameliorate Gut Inflammation via Toll-like Receptor 3 and Toll-like Receptor 7-Mediated Interferon-beta Production. Immunity 44, 889–900, doi:10.1016/j.immuni.2016.03.009 (2016).

20. Norman, J. M. et al. Disease-specific alterations in the enteric virome in inflammatory bowel disease. Cell 160, 447–460, doi:10.1016/j.cell.2015.01.002 (2015).

21. Cadwell, K. et al. Virus-plus-susceptibility gene interaction determines Crohn’s disease gene Atg16L1 phenotypes in intestine. Cell 141, 1135–1145, doi:10.1016/j.cell.2010.05.009 (2010).

22. Liang, G. et al. Dynamics of the Stool Virome in Very Early-Onset Inflammatory Bowel Disease. J Crohns Colitis 14, 1600–1610, doi:10.1093/ecco-jcc/jjaa094 (2020).

23. Wang, W. et al. Metagenomic analysis of microbiome in colon tissue from subjects with inflammatory bowel diseases reveals interplay of viruses and bacteria. Inflamm Bowel Dis 21, 1419–1427, doi:10.1097/MIB.0000000000000344 (2015).

24. Sartor, R. B. & Wu, G. D. Roles for Intestinal Bacteria, Viruses, and Fungi in Pathogenesis of Inflammatory Bowel Diseases and Therapeutic Approaches. Gastroenterology 152, 327–339 e324, doi:10.1053/j.gastro.2016.10.012 (2017).

25. Tarris, G. et al. Enteric Viruses and Inflammatory Bowel Disease. Viruses 13, doi:10.3390/v13010104 (2021).

26. Baldridge, M. T. et al. Commensal microbes and interferon-lambda determine persistence of enteric murine norovirus infection. Science 347, 266–269, doi:10.1126/science.1258025 (2015).

27. Pott, J. et al. IFN-lambda determines the intestinal epithelial antiviral host defense. Proc Natl Acad Sci U S A 108, 7944–7949, doi:10.1073/pnas.1100552108 (2011).

28. Mahlakoiv, T., Hernandez, P., Gronke, K., Diefenbach, A. & Staeheli, P. Leukocyte-derived IFN-alpha/beta and epithelial IFN-lambda constitute a compartmentalized mucosal defense system that restricts enteric virus infections. PLoS Pathog 11, e1004782, doi:10.1371/journal.ppat.1004782 (2015).

29. Lin, J. D. et al. Distinct Roles of Type I and Type III Interferons in Intestinal Immunity to Homologous and Heterologous Rotavirus Infections. PLoS Pathog 12, e1005600, doi:10.1371/journal.ppat.1005600 (2016).

30. Hernandez, P. P. et al. Interferon-lambda and interleukin 22 act synergistically for the induction of interferon-stimulated genes and control of rotavirus infection. Nat Immunol 16, 698–707, doi:10.1038/ni.3180 (2015).

31. Nice, T. J. et al. Interferon-lambda cures persistent murine norovirus infection in the absence of adaptive immunity. Science 347, 269–273, doi:10.1126/science.1258100 (2015).

32. Schneider, W. M., Chevillotte, M. D. & Rice, C. M. Interferon-stimulated genes: a complex web of host defenses. Annu Rev Immunol 32, 513–545, doi:10.1146/annurev-immunol-032713-120231 (2014).

33. Van Winkle, J. A. et al. Homeostatic interferon-lambda response to bacterial microbiota stimulates preemptive antiviral defense within discrete pockets of intestinal epithelium. Elife 11, doi:10.7554/eLife.74072 (2022).

34. Wirusanti, N. I., Baldridge, M. T. & Harris, V. C. Microbiota regulation of viral infections through interferon signaling. Trends Microbiol, doi:10.1016/j.tim.2022.01.007 (2022).

35. Kotenko, S. V. et al. IFN-lambdas mediate antiviral protection through a distinct class II cytokine receptor complex. Nat Immunol 4, 69–77, doi:10.1038/ni875 (2003).

36. Sheppard, P. et al. IL-28, IL-29 and their class II cytokine receptor IL-28R. Nat Immunol 4, 63–68, doi:10.1038/ni873 (2003).

37. Prokunina-Olsson, L. et al. A variant upstream of IFNL3 (IL28B) creating a new interferon gene IFNL4 is associated with impaired clearance of hepatitis C virus. Nat Genet 45, 164–171, doi:10.1038/ng.2521 (2013).

38. Katakura, K. et al. Toll-like receptor 9-induced type I IFN protects mice from experimental colitis. J Clin Invest 115, 695–702, doi:10.1172/JCI22996 (2005).

39. Pena Rossi, C. et al. Interferon beta-1a for the maintenance of remission in patients with Crohn’s disease: results of a phase II dose-finding study. BMC Gastroenterol 9, 22, doi:10.1186/1471-230X-9-22 (2009).

40. Pena-Rossi, C. et al. Clinical trial: a multicentre, randomized, double-blind, placebo-controlled, dose-finding, phase II study of subcutaneous interferon-beta-la in moderately active ulcerative colitis. Aliment Pharmacol Ther 28, 758–767 (2008).

41. Watanabe, T. et al. A case of exacerbation of ulcerative colitis induced by combination therapy with PEG-interferon alpha-2b and ribavirin. Gut 55, 1682–1683, doi:10.1136/gut.2006.105197 (2006).

42. Rauch, I. et al. Type I interferons have opposing effects during the emergence and recovery phases of colitis. Eur J Immunol 44, 2749–2760, doi:10.1002/eji.201344401 (2014).

43. Fan, J. B. et al. Type I IFN induces protein ISGylation to enhance cytokine expression and augments colonic inflammation. Proc Natl Acad Sci U S A 112, 14313–14318, doi:10.1073/pnas.1505690112 (2015).

44. Broggi, A., Tan, Y., Granucci, F. & Zanoni, I. IFN-lambda suppresses intestinal inflammation by non-translational regulation of neutrophil function. Nat Immunol 18, 1084–1093, doi:10.1038/ni.3821 (2017).

45. McElrath, C. et al. Critical role of interferons in gastrointestinal injury repair. Nat Commun 12, 2624, doi:10.1038/s41467-021-22928-0 (2021).

46. Chiriac, M. T. et al. Activation of Epithelial Signal Transducer and Activator of Transcription 1 by Interleukin 28 Controls Mucosal Healing in Mice With Colitis and Is Increased in Mucosa of Patients With Inflammatory Bowel Disease. Gastroenterology 153, 123–138 e128, doi:10.1053/j.gastro.2017.03.015 (2017).

47. Gunther, C. et al. Interferon Lambda Promotes Paneth Cell Death Via STAT1 Signaling in Mice and Is Increased in Inflamed Ileal Tissues of Patients With Crohn’s Disease. Gastroenterology 157, 1310–1322 e1313, doi:10.1053/j.gastro.2019.07.031 (2019).

48. Broggi, A., Granucci, F. & Zanoni, I. Type III interferons: Balancing tissue tolerance and resistance to pathogen invasion. J Exp Med 217, doi:10.1084/jem.20190295 (2020).

49. Odendall, C. et al. Diverse intracellular pathogens activate type III interferon expression from peroxisomes. Nat Immunol 15, 717–726, doi:10.1038/ni.2915 (2014).

50. Kotenko, S. V. IFN-lambdas. Curr Opin Immunol 23, 583–590, doi:10.1016/j.coi.2011.07.007 (2011).

51. Odendall, C. & Kagan, J. C. The unique regulation and functions of type III interferons in antiviral immunity. Curr Opin Virol 12, 47–52, doi:10.1016/j.coviro.2015.02.003 (2015).

52. Shouval, D. S. et al. Interleukin-10 receptor signaling in innate immune cells regulates mucosal immune tolerance and anti-inflammatory macrophage function. Immunity 40, 706–719, doi:10.1016/j.immuni.2014.03.011 (2014).

53. Forero, A. et al. Differential Activation of the Transcription Factor IRF1 Underlies the Distinct Immune Responses Elicited by Type I and Type III Interferons. Immunity 51, 451–464 e456, doi:10.1016/j.immuni.2019.07.007 (2019).

54. Anderson, C. A. et al. Meta-analysis identifies 29 additional ulcerative colitis risk loci, increasing the number of confirmed associations to 47. Nat Genet 43, 246–252, doi:10.1038/ng.764 (2011).

55. Zhang, J. X., Song, J., Wang, J. & Dong, W. G. JAK2 rs10758669 polymorphisms and susceptibility to ulcerative colitis and Crohn’s disease: a meta-analysis. Inflammation 37, 793–800, doi:10.1007/s10753-013-9798-5 (2014).

56. Franke, A. et al. Genome-wide meta-analysis increases to 71 the number of confirmed Crohn’s disease susceptibility loci. Nat Genet 42, 1118–1125, doi:10.1038/ng.717 (2010).

57. Lazear, H. M., Nice, T. J. & Diamond, M. S. Interferon-lambda: Immune Functions at Barrier Surfaces and Beyond. Immunity 43, 15–28, doi:10.1016/j.immuni.2015.07.001 (2015).

58. Clarke, L. et al. The international Genome sample resource (IGSR): A worldwide collection of genome variation incorporating the 1000 Genomes Project data. Nucleic Acids Res 45, D854–D859, doi:10.1093/nar/gkw829 (2017).

59. Karczewski, K. J. et al. The ExAC browser: displaying reference data information from over 60 000 exomes. Nucleic Acids Res 45, D840–D845, doi:10.1093/nar/gkw971 (2017).

60. Mendoza, J. L. et al. The IFN-lambda-IFN-lambdaR1-IL-10Rbeta Complex Reveals Structural Features Underlying Type III IFN Functional Plasticity. Immunity 46, 379–392, doi:10.1016/j.immuni.2017.02.017 (2017).

61. Fox, B. A., Sheppard, P. O. & O’Hara, P. J. The role of genomic data in the discovery, annotation and evolutionary interpretation of the interferon-lambda family. PLoS One 4, e4933, doi:10.1371/journal.pone.0004933 (2009).

62. McCracken, K. W., Howell, J. C., Wells, J. M. & Spence, J. R. Generating human intestinal tissue from pluripotent stem cells in vitro. Nat Protoc 6, 1920–1928, doi:10.1038/nprot.2011.410 (2011).

63. Hadjadj, J. et al. Impaired type I interferon activity and inflammatory responses in severe COVID-19 patients. Science 369, 718–724, doi:10.1126/science.abc6027 (2020).

64. Haga, K. et al. Genetic Manipulation of Human Intestinal Enteroids Demonstrates the Necessity of a Functional Fucosyltransferase 2 Gene for Secretor-Dependent Human Norovirus Infection. mBio 11, doi:10.1128/mBio.00251-20 (2020).

65. Peterson, S. T. et al. Disruption of Type III Interferon (IFN) Genes Ifnl2 and Ifnl3 Recapitulates Loss of the Type III IFN Receptor in the Mucosal Antiviral Response. J Virol 93, doi:10.1128/JVI.01073-19 (2019).

66. Hong, M. et al. Interferon lambda 4 expression is suppressed by the host during viral infection. J Exp Med 213, 2539–2552, doi:10.1084/jem.20160437 (2016).

67. Knapp, S., Meghjee, N., Cassidy, S., Jamil, K. & Thursz, M. Detection of allele specific differences in IFNL3 (IL28B) mRNA expression. BMC Med Genet 15, 104, doi:10.1186/s12881-014-0104-7 (2014).

68. Sposito, B. et al. The interferon landscape along the respiratory tract impacts the severity of COVID-19. Cell, doi:10.1016/j.cell.2021.08.016 (2021).

69. Van Winkle, J. A. et al. A homeostatic interferon-lambda response to bacterial microbiota stimulates preemptive antiviral defense within discrete pockets of intestinal epithelium. Elife 11, doi:10.7554/eLife.74072 (2022).

70. Vardi, I. et al. Genetic and Structural Analysis of a SKIV2L Mutation Causing Tricho-hepato-enteric Syndrome. Dig Dis Sci 63, 1192–1199, doi:10.1007/s10620-018-4983-x (2018).

71. Li, H. & Durbin, R. Fast and accurate short read alignment with Burrows-Wheeler transform. Bioinformatics 25, 1754–1760, doi:10.1093/bioinformatics/btp324 (2009).

72. McKenna, A. et al. The Genome Analysis Toolkit: a MapReduce framework for analyzing next-generation DNA sequencing data. Genome Res 20, 1297–1303, doi:10.1101/gr.107524.110 (2010).

73. Li, M. X., Gui, H. S., Kwan, J. S., Bao, S. Y. & Sham, P. C. A comprehensive framework for prioritizing variants in exome sequencing studies of Mendelian diseases. Nucleic Acids Res 40, e53, doi:10.1093/nar/gkr1257 (2012).

74. Lyons, J. J. et al. Elevated basal serum tryptase identifies a multisystem disorder associated with increased TPSAB1 copy number. Nat Genet 48, 1564–1569, doi:10.1038/ng.3696 (2016).

75. Park, S. & Mostoslavsky, G. Generation of Human Induced Pluripotent Stem Cells Using a Defined, Feeder-Free Reprogramming System. Curr Protoc Stem Cell Biol 45, e48, doi:10.1002/cpsc.48 (2018).

76. Watson, C. L. et al. An in vivo model of human small intestine using pluripotent stem cells. Nature medicine 20, 1310–1314, doi:10.1038/nm.3737 (2014).

77. Sato, T. et al. Long-term expansion of epithelial organoids from human colon, adenoma, adenocarcinoma, and Barrett’s epithelium. Gastroenterology 141, 1762–1772, doi:10.1053/j.gastro.2011.07.050 (2011).

