## Supplemental Figure 1 for "Variants in Interferon Lambda are Associated with Very Early Onset Inflammatory Bowel Disease"

**Supplemental Figures:**

**Supplemental Figure 1: Dose response curves in response to WT and patient-encoded variants.**

**(A)** EC50 comparison between IFN-λ2 and IFN-λ2Var from dose response curve described in Figure 2A.

**(B)** Dose-response curve of downstream ISG induction following stimulation with either IFN-λ3 or IFN-λ3Var1. Normalized relative luciferase units (RLU) shown.

**(C)** EC50 comparison between IFN-λ3 and IFN-λ3Var1. EC50 not quantifiable for IFN-λ3Var1.

**(D)** Dose-response curve of downstream ISG induction following stimulation with either IFN-λ3 or IFN-λ3Var2. Normalized relative luciferase units (RLU) shown.

**(E)** EC50 comparison between IFN-λ3 and IFN-λ3Var2.

**(F)** Induction of ISGs following stimulation of healthy control tHIOs with either WT IFN-λ2 (yellow dots), patient-encoded IFN-λ2Var (peach dots), or no treatment (black dots). HP1 = Healthy parent 1; HP2 = healthy parent 2; SI = small intestinal enteroids of unrelated healthy control

**(G)** EC50 comparison between INF-λ2 and INF-λ2Var in dimerization assay with IFNLR1 and IL10RB

(**H**) EC50 comparison between INF-λ3 and INF-λ3Var1 in dimerization assay with IFNLR1 and IL10RB

(B-E, G, H) Data depicts three independent experiments, each data point is an independent experiment. RLU were normalized to blank wells (no protein). (C,E,G,H) Statistical significance determined by unpaired t-test between the groups.

### Supplemental Figure 1

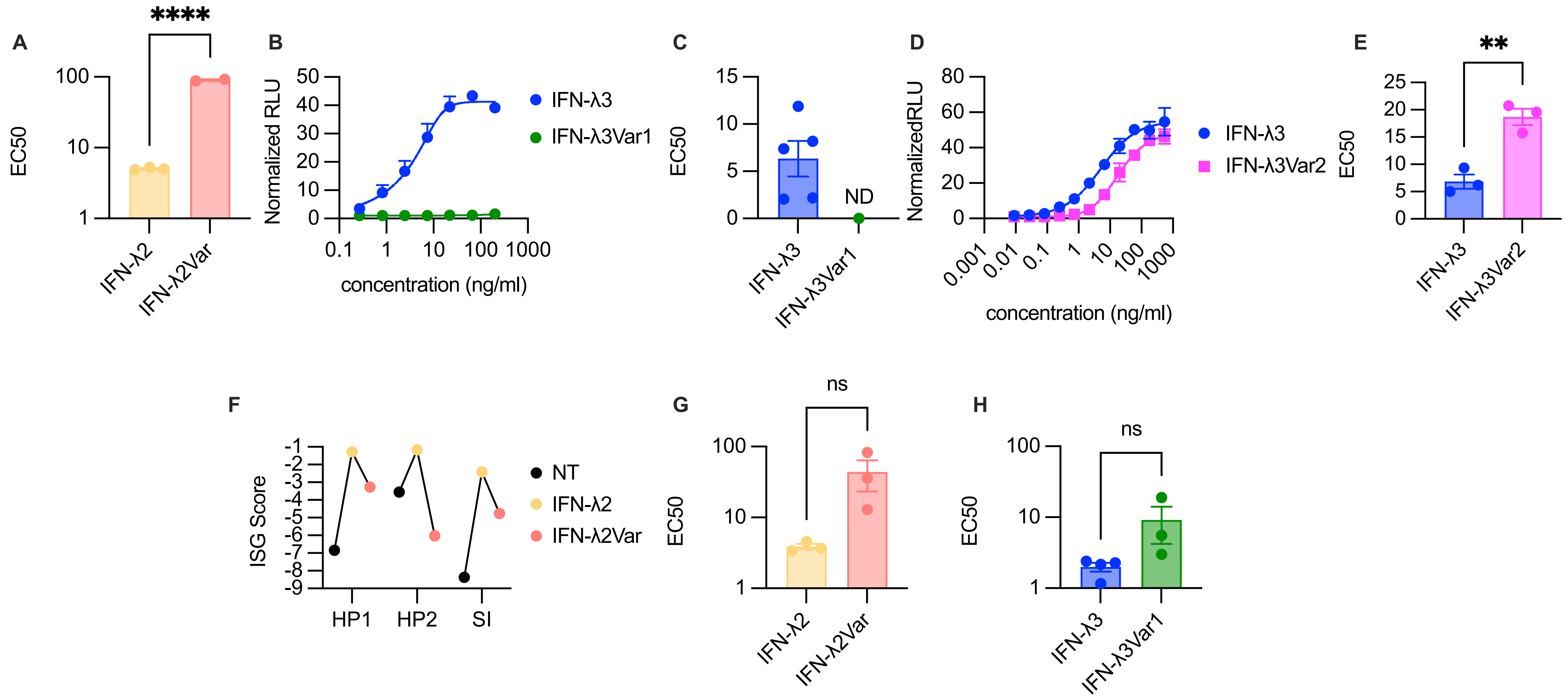
